# Association of free fatty acids with long-term adverse outcomes in patients with premature myocardial infarction: a prospective cohort study

**DOI:** 10.1101/2025.04.08.25325497

**Authors:** Jing Wang, Jing-xian Wang, Saidinisa Ruzemaimaiti, Yu-hang Wang, Yu Zhou, Jing-xi Chen, Chang-ping Li, Zhuang Cui, Yin Liu, Jing Gao

## Abstract

**Background:** The dose-response relationship and independent prognostic value of free fatty acids (FFA) on major adverse cardiovascular events (MACE) in patients with premature myocardial infarction (PMI) are not well defined. This study aims to explore the significance of FFA levels on long-term prognostic in patients with PMI and the combined effects of FFA with inflammatory markers, obesity, and insulin resistance (IR) on clinical outcomes.

**Methods:** This single-center prospective cohort study enrolled 1,168 consecutive patients with PMI admitted to the CCU ward of Tianjin Chest Hospital from March 2017 to December 2024. Participants were stratified into four groups (Q1-Q4) based on the quartiles of baseline FFA levels. With a median follow-up of 2.83 years (IQR: 2.58–3.10), the primary endpoint was MACE. Cox proportional hazard models assessed the association between FFA levels and long-term outcomes, while the restricted cubic spline plot evaluated the nonlinear relationship between FFA levels and MACE risk.

**Results:** 216 (18.50%) MACE were recorded during a median follow-up of 2.83 years. FFA levels showed a non-linear U-shaped association with long-term prognosis in patients with PMI. When the second quartile of FFA levels (Q2: 0.43-0.61 mmol/L) was used as the reference group, after multifactorial correction, the association between FFA levels in the Q1 (<0.43 mmol/L) [HR=1.888 (95% CI: 1.109-2.956), *p*=0.009], Q3 (0.61-0.83 mmol/L) [HR=2.466 (95% CI: 1.548-3.928), *p*<0.001] and Q4 (≥0.83 mmol/L) [HR=2.944 (95% CI: 1.862-4.654), *p*<0.001] had significantly higher risk of MACE. Subgroup analyses showed that the U-shaped association between FFA and MACE remained significant in PMI patients with comorbid metabolic syndrome or type 2 diabetes mellitus. Subsequent joint analyses demonstrated that patients with PMI exhibited a significantly elevated risk of MACE if they had abnormal levels of FFA and a metabolic disorder with a hypersensitive C-reactive protein (hs-CRP) >5 mg/L, body mass index (BMI) >28 kg/m², or TyG >9.21 mmol/L.

**Conclusions:** Elevated and decreased levels of FFA, when surpassing a certain threshold, independently increase the risk of MACE in patients with PMI. Consequently, dynamic monitoring of FFA and combined assessment of inflammation, obesity, and IR in patients with PMI are essential for risk stratification and clinical management.

## Introduction

The epidemiological characteristics of acute myocardial infarction (AMI) demonstrate a pronounced tendency towards younger demographics^1^, which has emerged as a salient challenge in global public health ^2,3^. There is an urgent need for more effective strategies to facilitate rapid diagnosis, risk stratification, and management to improve long-term prognosis in PMI populations.

Free fatty acids (FFA) are a product of triglyceride catabolism and serve as a significant energy source during human metabolism. Basic studies have demonstrated that elevated FFA levels can contribute to the process of atherosclerotic plaque formation and destabilization by inducing vascular endothelial dysfunction^4^, exacerbating insulin resistance ^5,6^, triggering the oxidative stress cascade^7,8^, promoting ectopic lipid deposition, and mediating lipotoxicity damage, among other molecular pathways. Meanwhile, several studies have demonstrated that elevated levels of FFA can predict total and cardiovascular mortality in various populations ^9,10^. A cohort study from 26 countries demonstrated that FFA levels were directly associated with the risk of MACE and death in patients with type 2 diabetes mellitus (T2DM) combined with recent acute coronary syndrome (ACS)^11^. The PRACTICE study corroborated a nonlinear U-shaped correlation between baseline FFA levels and mortality or ischemic events in patients with coronary artery disease and T2DM^12^. In patients with PMI, the characteristics of coronary artery lesions and the risk factors differ from those observed in traditional cardiovascular disease populations^13,14^. Combination of multiple metabolic disorders with predominantly glycolipid metabolism disorders in the PMI population. Previous research conducted by our team has demonstrated that comorbid metabolic syndrome (MS) has been identified as a significant independent risk factor for MACE in the PMI population, that hyperglycemia in the MS component often predicted more severe coronary heart disease, and that the MS component (including BMI≥28.0 kg/m^2^ and hyperglycemia) was independently associated with the development of MACE^15^. Concentrations of FFA have been identified as a modifiable risk factor for metabolic diseases and may serve as a novel biological target for the prevention and treatment of cardiovascular diseases^16^. However, a paucity of large prospective studies exists that examine the relationship between FFA levels and long-term prognosis in patients with PMI. In the present study, therefore, we sought to analyze the dose-effect relationship between FFA levels and long-term MACE, as well as its independent predictive value. We also sought to explore the synergistic effect of FFA levels with metabolic disorders such as high inflammation, obesity, and IR on clinical prognosis.

## Methods

### Study design and participants

This study included 1280 consecutive patients with PMI treated with percutaneous coronary intervention (PCI) in the CCU ward of Tianjin Chest Hospital between March 2017 and December 2024. Based on the established exclusion criteria, 112 patients were excluded, and the final sample consisted of 1168 patients. The study flow chart is presented in Figure1.

The inclusion criteria, as outlined in the fourth edition of the global harmonized definition of myocardial infarction, are as follows: elevated and/or decreased cTn, exceeding the 99th percentile of the upper limit of normal at least once, and at least one of the following: Firstly, symptoms of myocardial ischemia. Secondly, new ischemic electrocardiographic changes. Thirdly, new pathologic Q-waves. Fourthly, imaging suggests deletion of new surviving myocardium consistent with ischemia or segmental ventricular motion abnormalities. Fifthly, coronary artery thrombosis is confirmed by coronary angiography or autopsy. Patients diagnosed with acute myocardial infarction based on the aforementioned criteria who underwent coronary angiography and whose age was consistent with ≤45 years for men and ≤55 years for women were included in this study. The exclusion criteria included the following: (1) an inability to obtain FFA data; (2) the presence of heart valve disease, rheumatic heart disease, aortic coarctation, pulmonary embolism, pulmonary heart disease, congenital heart disease, or severe heart failure; (3) the presence of malignant tumors with an expected survival time of less than 1 year; and (4) severe hepatic and renal dysfunction.

### Clinical and Biochemical Measurements

The data were retrieved from the electronic medical records and encompassed the following information: demographic and baseline characteristics (sex, age, BMI); cardiovascular risk factors (smoking and alcohol history); cardiovascular history, including previous PCI or coronary artery bypass grafting (CABG) procedures; family history of premature coronary artery disease; admission vital signs (heart rate and blood pressure); and clinical status (Killip classification and type of AMI). The laboratory parameters encompassed a range of indicators of cardiac injury, including cardiac troponin T (cTnT) and creatine kinase MB (CK-MB), as well as parameters of glucose and lipid metabolism, such as fasting plasma glucose (FPG), total cholesterol (TC), triglycerides (TG), high-density lipoprotein cholesterol (HDL-C) and low-density lipoprotein cholesterol (LDL-C). In addition, inflammatory markers, including high-sensitivity C-reactive protein (hs-CRP), were measured, as were hepatic and renal function parameters. Medication regimens included antiplatelet agents, angiotensin-converting enzyme inhibitors (ACEI) or angiotensin receptor blockers (ARB), statins, β-blockers, and others. Interventional data covered the severity of coronary stenosis, stent characteristics (location, number, size), and the performance of the procedure by certified interventional cardiologists.

### Free fatty acid measurement

Patients were required to fast for 4-12 hours after admission, after which fasting blood samples were collected. The blood samples were subjected to centrifugation at 4°C, 3000 rpm for 10 minutes, and the separated serum was frozen and stored at -80°C. Serum FFA levels were determined by the standard enzyme colorimetric method of the Department of Clinical Laboratory, Tianjin Chest Hospital, with a reference range of 0.1-0.6 mmol/L. The study population was then categorized based on the quartiles of baseline FFA levels, resulting in the division of the population into four groups (Q1: <0.43 mmol/L, Q2: 0.43-0.61 mmol/L, Q3: 0.61-0.83 mmol/L, Q4: ≥0.83 mmol/L).

### Follow-Up and Outcomes

The median follow-up duration was 2.83 years (IQR: 2.58–3.10), and patients were continuously monitored by trained investigators through outpatient visits, telephone contacts, and questionnaires. Patients were assessed for medication adherence and adverse events during follow-up. The study endpoints were MACE, including cardiac death, rehospitalization for severe heart failure, non-fatal myocardial infarction, ischemic stroke, readmission for unstable angina, and target lesion revascularization (TLR), defined as unplanned revascularization driven by an ischemic symptom or event, including PCI and CABG.

### Statistical Analysis

The statistical analysis of the study was conducted using SPSS 25.0 and R 4.4.1 software. The Kolmogorov-Smirnov test was employed to assess the normality of the data. Variables that exhibited a normal distribution were described as mean ± standard deviation, and comparisons between groups were made using either an independent samples t-test (for two groups) or a one-way analysis of variance (for multiple groups). Variables that did not exhibit a normal distribution were expressed as median (interquartile range [IQR]), and comparisons between groups were made using the Mann-Whitney U test (for two groups) or the Kruskal-Wallis test (for multiple groups); categorical variables were expressed as frequencies (percentages), and differences between groups were assessed by the chi-square test or Fisher’s exact method (for theoretical frequencies < 1 or sample sizes < 40). Event-free survival was analyzed using Kaplan-Meier curves, and Log-rank tests were used to compare differences between groups. To calculate risk ratios (HR, 95% CI) for adverse outcomes, we employed one-way and multifactor Cox regression. We resolved the nonlinear association between FFA and MACE using a restricted cubic spline (RCS) plot. We defined statistical significance as a two-sided p-value of < 0.05.

## Results

### Baseline characteristics

The mean age of the 1168 patients with PMI included in this study was 41.01 ± 5.77 years, of which 1017 (87.10%) were male. The study population was categorized into four groups based on quartiles of FFA levels, and the baseline characteristics of each group based on FFA levels are detailed in Table1. The proportion of male patients was lower among those with high FFA levels (groups Q3 and Q4) compared to those with low FFA levels (groups Q1 and Q2). BMI, blood pressure, inflammation markers, liver function indicators, glucose metabolism indicators, lipid metabolism indicators, and myocardial injury markers were significantly higher in patients with high FFA levels than in those with low FFA levels. The type of myocardial infarction was associated with FFA levels, with a higher proportion of patients experiencing ST-segment Elevation Myocardial Infarction (STEMI) among those with high FFA levels.

### Association of FFA with MACE in PMI patients

During a median follow-up period of 2.83 years (IQR: 2.58–3.10), a total of 216 (18.50%) MACEs were recorded, including 15 cardiac deaths, 4 ischemic strokes, 13 nonfatal myocardial infarctions, 44 readmissions for severe heart failure, 86 target lesion revascularizations and 54 readmissions for unstable angina. The baseline characteristics of the PMI population according to the occurrence of MACE are detailed in Table 2. The levels of FPG, glycated hemoglobin (HbA1c), TyG index, hs-CRP, N-terminal pro-brain natriuretic peptide (NT-ProBNP), and FFA were significantly higher than in patients without MACE.

**Table 1:**
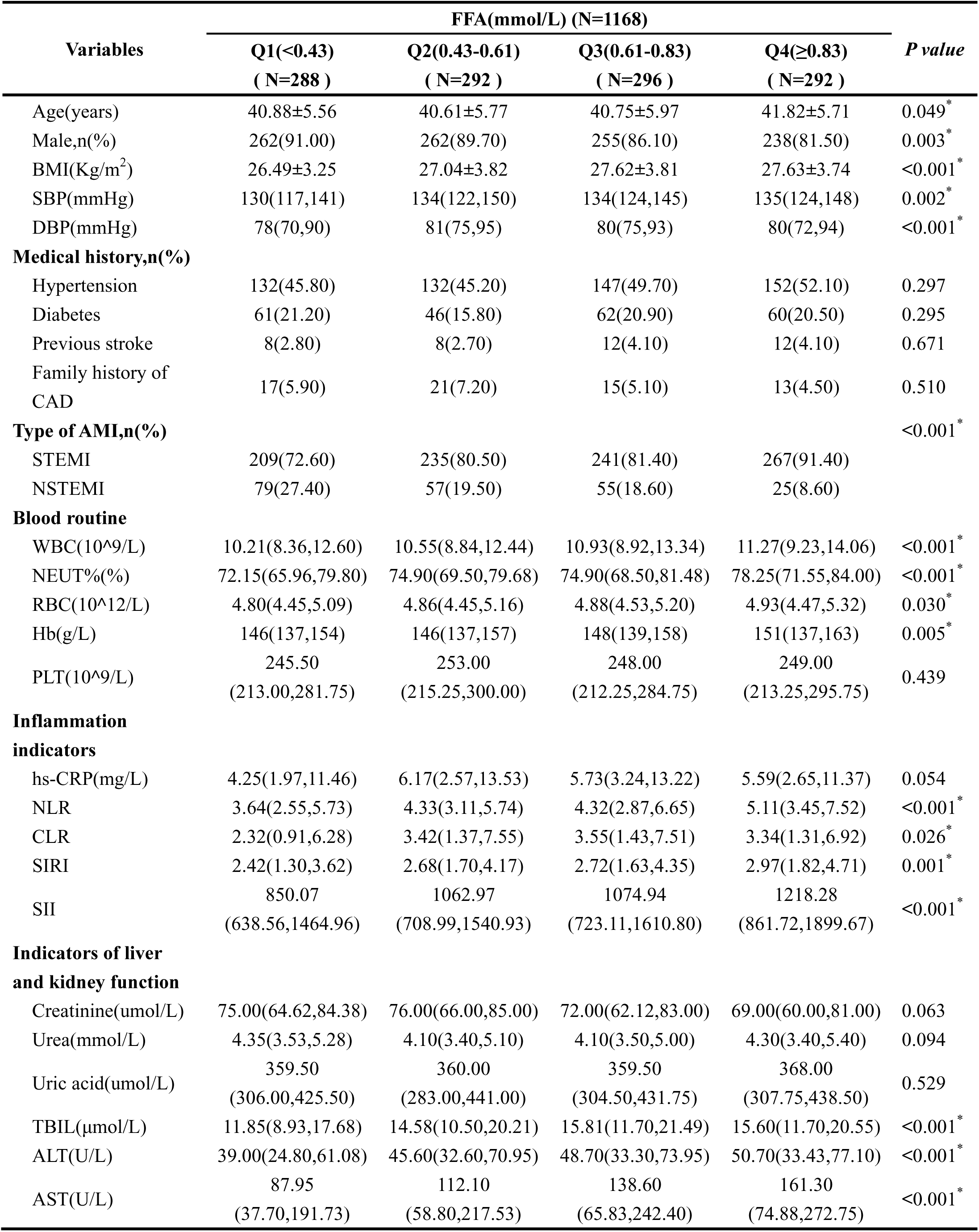
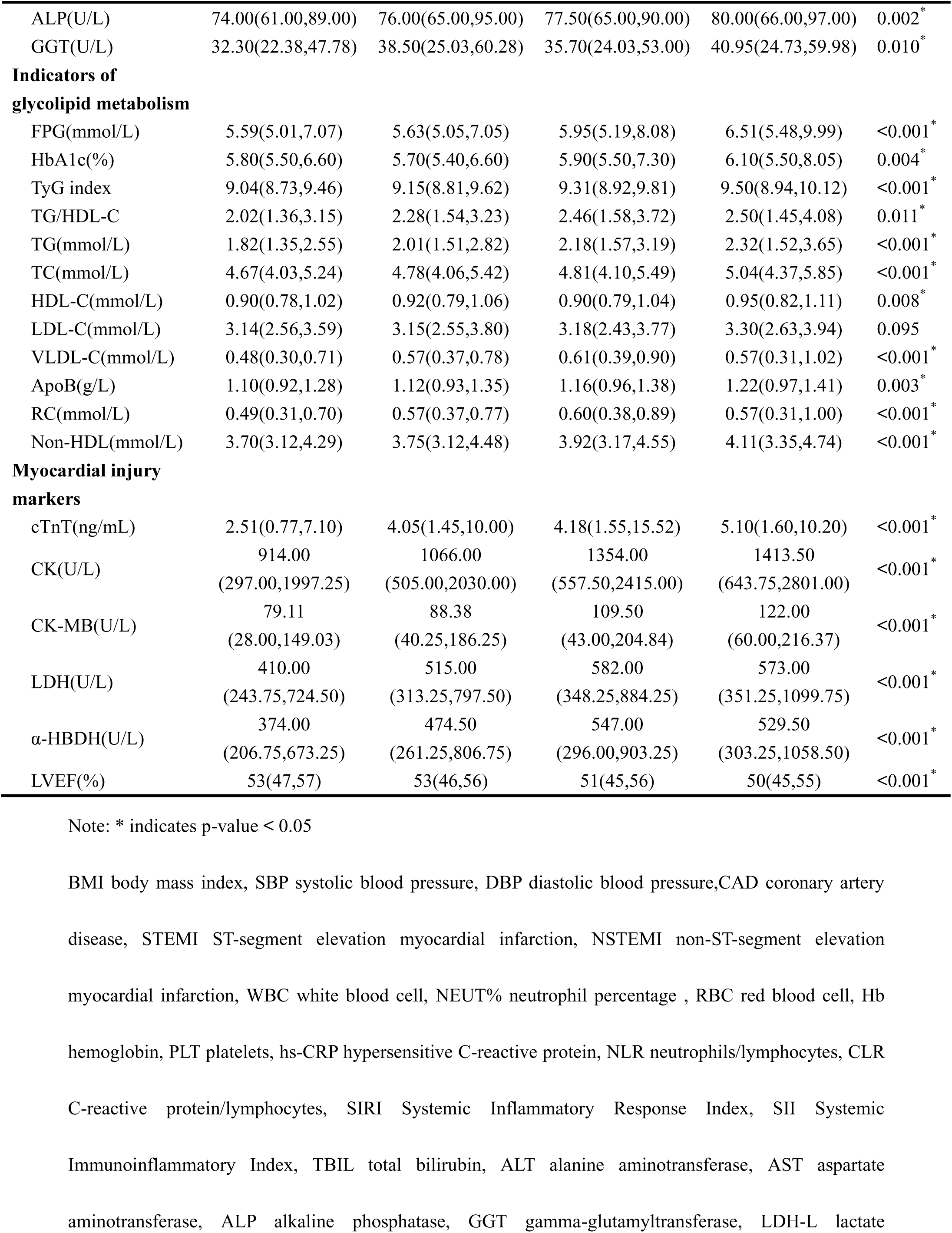
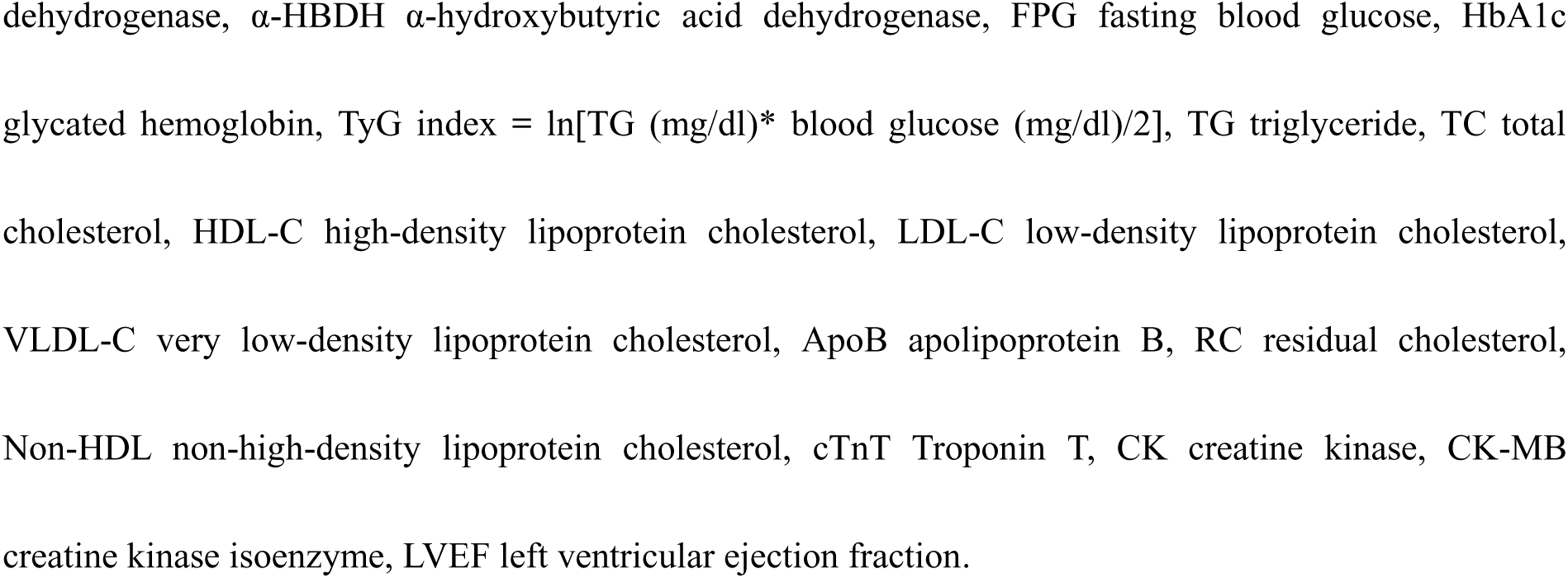
Baseline characteristics of all patients with PMI in different FFA levels.

**Table 2:** Baseline characteristics of the patients stratified by MACE.

| Variables | Total<br>( N=1168 ) | Non-MACE<br>( N=952 ) | MACE<br>(N=216) | *P value* |
| --- | --- | --- | --- | --- |
| Age(years) | 41.01±5.77 | 40.89±5.74 | 41.56±5.84 | 0.131 |
| Male,n(%) | 1017(87.10) | 832(87.40) | 185(85.60) | 0.490 |
| BMI(Kg/m<sup>2</sup>) | 27.20±3.69 | 27.13±3.72 | 27.53±3.51 | 0.138 |
| SBP(mmHg) | 134(121,145) | 133(121,145) | 135(122,150) | 0.383 |
| DBP(mmHg) | 80(72,92) | 80(72,91) | 81(71,95) | 0.762 |
| **Medical history,n(%)** |  |  |  |  |
| Hypertension | 563(48.20) | 449(47.20) | 114(52.80) | 0.136 |
| Diabetes | 229(19.60) | 168(17.60) | 61(28.20) | <0.001* |
| Previous stroke | 40(3.40) | 33(3.50) | 7(3.20) | 0.869 |
| Family history of CAD | 66(5.70) | 53(5.60) | 13(6.00) | 0.795 |
| **Laboratory** |  |  |  |  |
| WBC(10<sup>9</sup>/L) | 10.78(8.86,13.02) | 10.78(8.88,13.02) | 10.74(8.84,13.11) | 0.805 |
| NEUT%(%) | 75.30(68.90,81.10) | 75.30(69.00,81.10) | 75.10(68.73,80.98) | 0.989 |
| hs-CRP(mg/L) | 5.55(2.56,12.20) | 5.19(2.44,11.64) | 6.63(2.84,15.53) | 0.012* |
| NLR | 4.42(2.96,6.42) | 4.44(2.95,6.37) | 4.27(2.99,6.52) | 0.775 |
| CLR | 3.18(1.26,7.18) | 3.01(1.24,7.00) | 3.91(1.56,8.51) | 0.007* |
| SIRI | 2.70(1.61,4.27) | 2.67(1.61,4.20) | 2.87(1.61,4.33) | 0.493 |
| SII | 1070.85<br>(704.19,1633.53) | 1175.23<br>(704.19,1616.08) | 1052.74<br>(695.20,1686.62) | 0.850 |
| Cr(umol/L) | 73.00(63.00,83.00) | 73.00(63.00,82.49) | 75.50(63.00,89.00) | 0.059 |
| UA(umol/L) | 362.00(300.50,432.75) | 361.00(302.00,431.00) | 363.50(298.00,442.50) | 0.755 |
| ALT (U/L) | 45.50(30.00,71.40) | 46.25(30.13,70.70) | 43.00(29.03,76.43) | 0.839 |
| AST(U/L) | 124.30(55.73,236.15) | 125.30(56.60,234.67) | 114.85(52.93,241.48) | 0.949 |
| FPG(mmol/L) | 5.96(5.16,7.77) | 5.84(5.11,7.35) | 6.77(5.48,10.21) | <0.001* |
| HbA1c(%) | 5.80(5.50,7.20) | 5.80(5.40,6.83) | 6.25(5.60,8.73) | <0.001* |
| TyG index | 9.21(8.82,9.77) | 9.19(8.79,9.72) | 9.37(8.94,10.06) | <0.001* |
| TG(mmol/L) | 2.06(1.47,3.09) | 2.05(1.46,3.06) | 2.12(1.50,3.18) | 0.363 |
| TC(mmol/L) | 4.81(4.11,5.48) | 4.80(4.11,5.46) | 4.84(4.09,5.60) | 0.729 |
| HDL-C(mmol/L) | 0.92(0.80,1.05) | 0.92(0.81,1.05) | 0.91(0.77,1.07) | 0.294 |
| LDL-C(mmol/L) | 3.18(2.54,3.78) | 3.19(2.56,3.77) | 3.14(2.44,3.93) | 0.696 |
| VLDL-C(mmol/L) | 0.55(0.34,0.82) | 0.56(0.34,0.82) | 0.53(0.36,0.84) | 0.994 |
| ApoB(g/L) | 1.14(0.94,1.37) | 1.14(0.94,1.37) | 1.14(0.96,1.38) | 0.824 |
| RC(mmol/L) | 0.55(0.34,0.81) | 0.56(0.34,0.81) | 0.54(0.36,0.84) | 0.734 |
| Non-HDL(mmol/L) | 3.86(3.16,4.55) | 3.84(3.16,4.53) | 3.96(3.16,4.60) | 0.574 |
| NT-ProBNP (pg/mL) | 89.34<br>(10.00,321.86) | 81.13<br>(8.05,274.45) | 150.29<br>(12.95,536.43) | <0.001* |
| cTnT(ng/mL) | 3.75(1.32,10.00) | 3.89(1.36,10.00) | 3.34(1.20,8.88) | 0.093 |
| CK-MB(U/L) | 100.00(41.00,192.82) | 102.00(42.00,192.20) | 95.50(38.00,195.25) | 0.372 |
| LVEF(%) | 51(45,56) | 52(46,56) | 50(44,56) | 0.027* |
| FFA((mmol/L) | 0.61(0.43,0.83) | 0.60(0.43,0.80) | 0.70(0.43,0.94) | 0.003* |

The Kaplan-Meier survival curves categorized according to FFA levels are shown in Figure 2(A-C). Among the four groups of patients with varying FFA levels, the population in the Q2 group exhibited the highest cumulative event-free survival. The discrepancy between groups Q2 and Q3 did not attain statistical significance (P=0.058). However, group Q4 had the lowest (Q4 vs. Q2, P<0.001). Subgroup analyses demonstrated that the aforementioned conclusions remained valid in PMI populations with comorbid MS and T2DM. When the second quartile of FFA levels was used as the reference group, after multifactorial correction, the association between FFA levels in the Q1 group [HR=1.888 (95% CI: 1.109-2.956), p=0.009], Q3 group [HR=2.466 (95% CI: 1.548-3.928), p<0.001], and Q4 group [HR=2.944 (95% CI: 1.862-4.654), p<0.001] indicated a significantly higher risk of MACE (see Table3). On the continuous scale, the RCS plot revealed a nonlinear U-shaped relationship between FFA levels and MACE in the PMI population (P-nonlinear=0.002). As illustrated in Figure 2(E and F), subgroup analyses showed that analogous outcomes were observed in PMI patients with comorbid MS (P-nonlinear<0.001) and T2DM (P-nonlinear=0.015).

**Figure 1:**
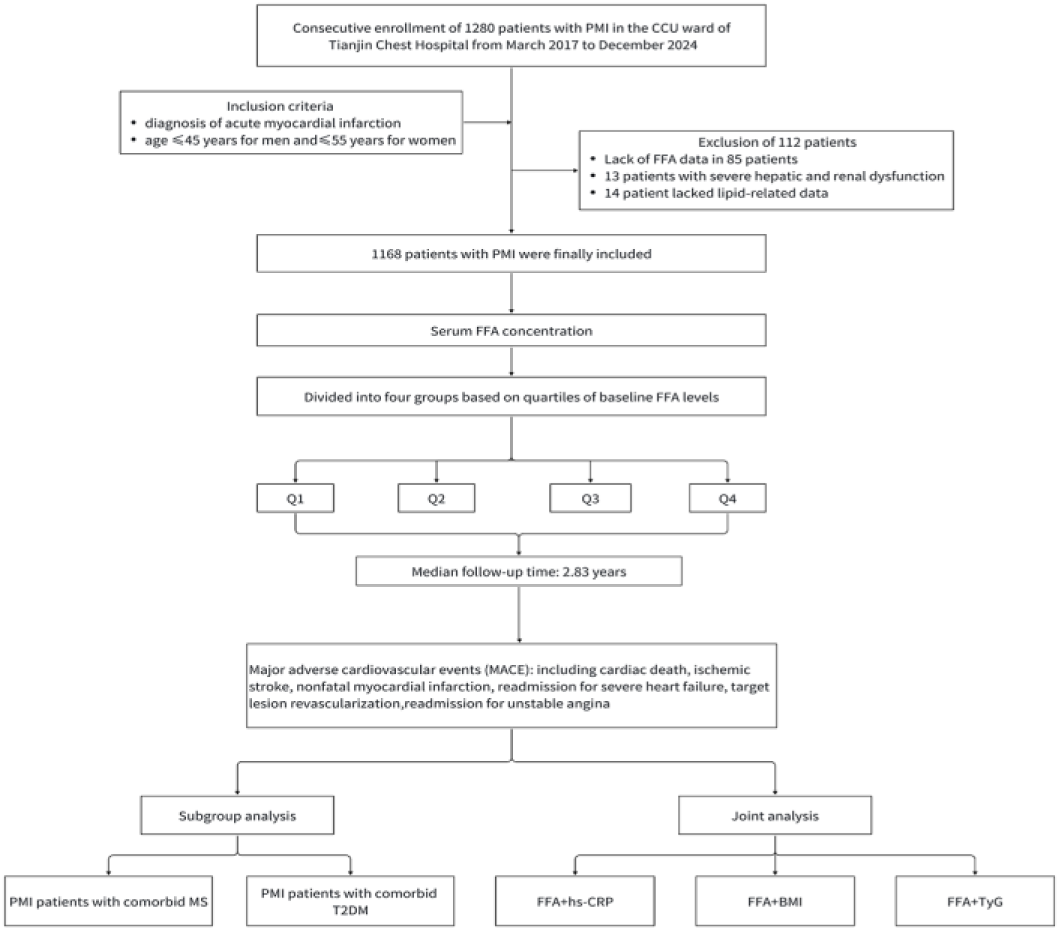
Research flow chart.

**Figure 2:**
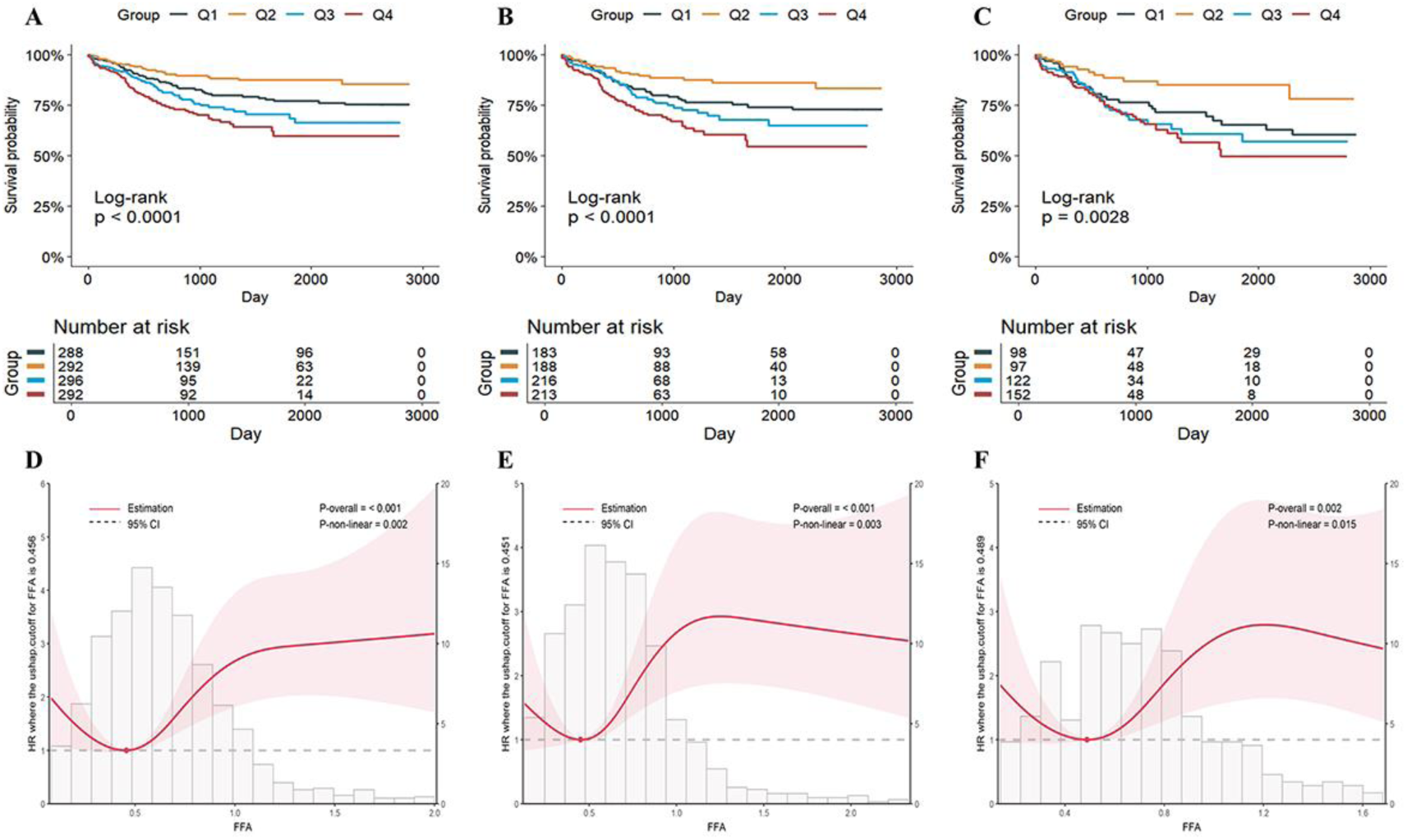
Kaplan-Meier curves and Restricted cubic spline plots for risk of MACE in PMI patients. **A-C** Kaplan-Meier curves for risk of MACE in PMI patients grouped according to FFA levels; **D-F** Restricted cubic spline (RCS) plots for risk of MACE according to FFA on a continuous scale; **A and D** Total PMI population; **B and E** PMI combined metabolic syndrome (MS) population; **C and F** PMI combined T2DM population.

To further explore the relationship between FFA levels and the risk of developing MACE in different ages, genders, and combined with MS or T2DM populations, the forest plot of subgroup analysis is shown in Figure 3. The results showed that the interaction between the above variables and FFA was not statistically significant and that FFA levels and MACE risk were not affected by sex, age, and comorbidities.

**Figure 3:**
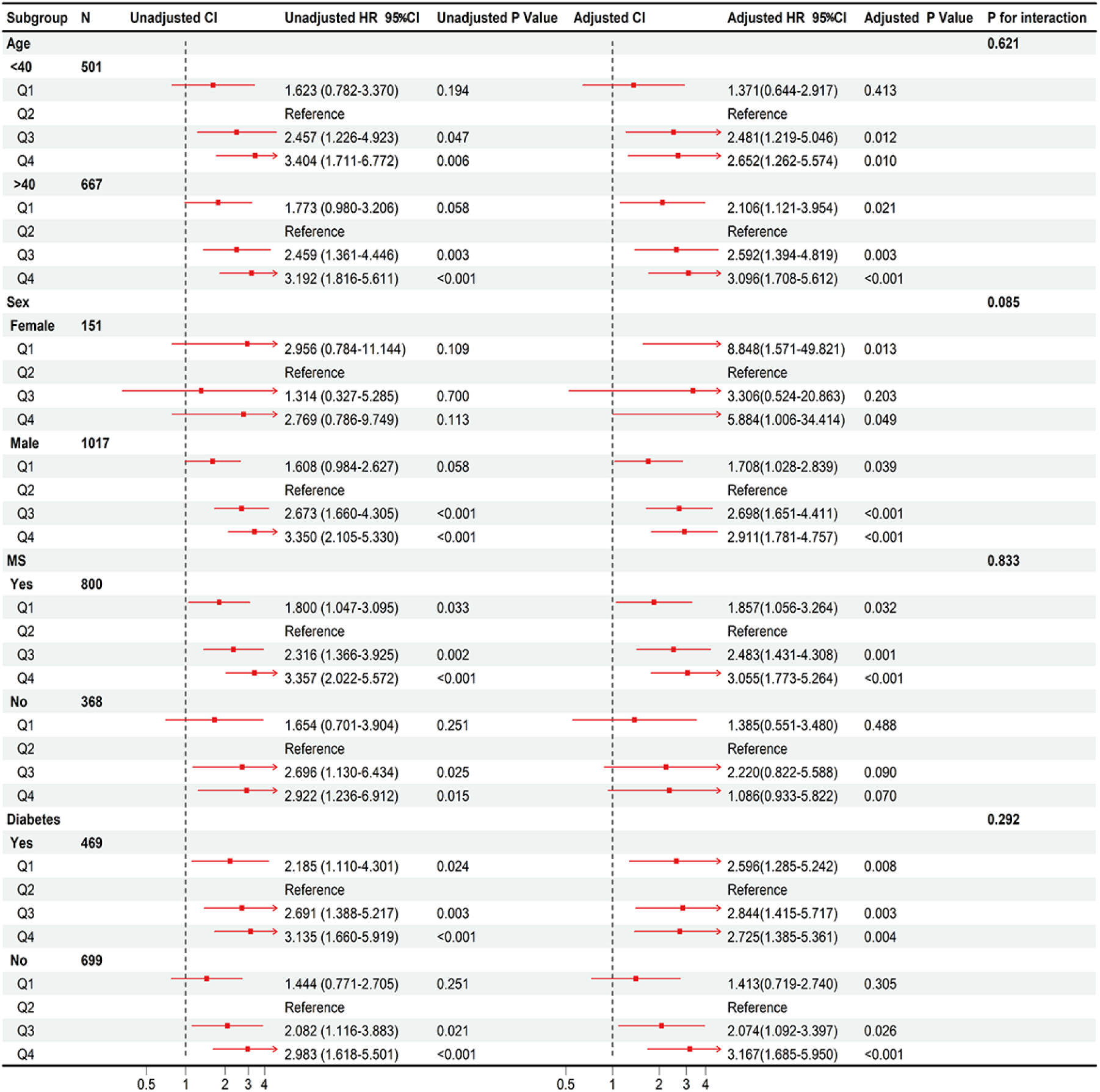
Subgroup analysis and forest plot according to sex, age, and comorbidities (MS, T2DM). Adjust: gender, age, BMI, heart rate, systolic blood pressure, diastolic blood pressure, history of smoking, history of alcohol consumption, history of hypertension, history of diabetes, history of PCI, and history of statin therapy, FPG, Cr, RC, D-Dimer, Comorbidity with metabolic syndrome.

### Effect of FFA levels in combination with hs-CRP, BMI, and TyG on MACE risk in a PMI population

The multifactorial Cox proportional hazards model demonstrated that elevated or reduced FFA levels were significantly associated with an elevated risk of MACE in PMI patients with a concomitant high-inflammatory state (hs-CRP >5 mg/L), obesity (BMI >28 kg/m²), or insulin resistance (TyG index >9.21mmol/L). Specifically, in the high-inflammation subgroup, the risk of poor prognosis was significantly higher when FFA was at Q1 [HR=2.480 (95% CI: 1.310-4.695], Q3 [HR=2.864 (95% CI: 1.548 -5.301], and Q4 [HR=3.051 (95% CI: 1.664-5.594] compared to the reference group (Q2 group) were significantly higher (respective *p-values* <0.05). These findings were consistent in the obesity and insulin resistance subgroups (see Table 3, Model 3). Specifically, Compared to the reference group (Q2 group), when BMI >28 kg/m², the risk of MACE escalated by 1.802, 2.824, and 2.499 fold, respectively, in the Q1, Q3, and Q4 groups. Furthermore, when the TyG index >9.21mmol/L, the risk of MACE increased 1.183, 1.594, and 2.282 fold for Q1, Q3, and Q4 groups, respectively. Patients with PMI were categorized into eight groups based on FFA levels and various metabolic disorder states, including inflammation levels, obesity, and insulin resistance. The long-term prognosis of these patients was then evaluated. The findings revealed that patients with levels of hs-CRP>5+Q4, BMI>28+Q4, and TyG>9.21+Q4 exhibited a significantly lower cumulative event-free survival compared to the control group (see Figure 4).

**Figure 4:**
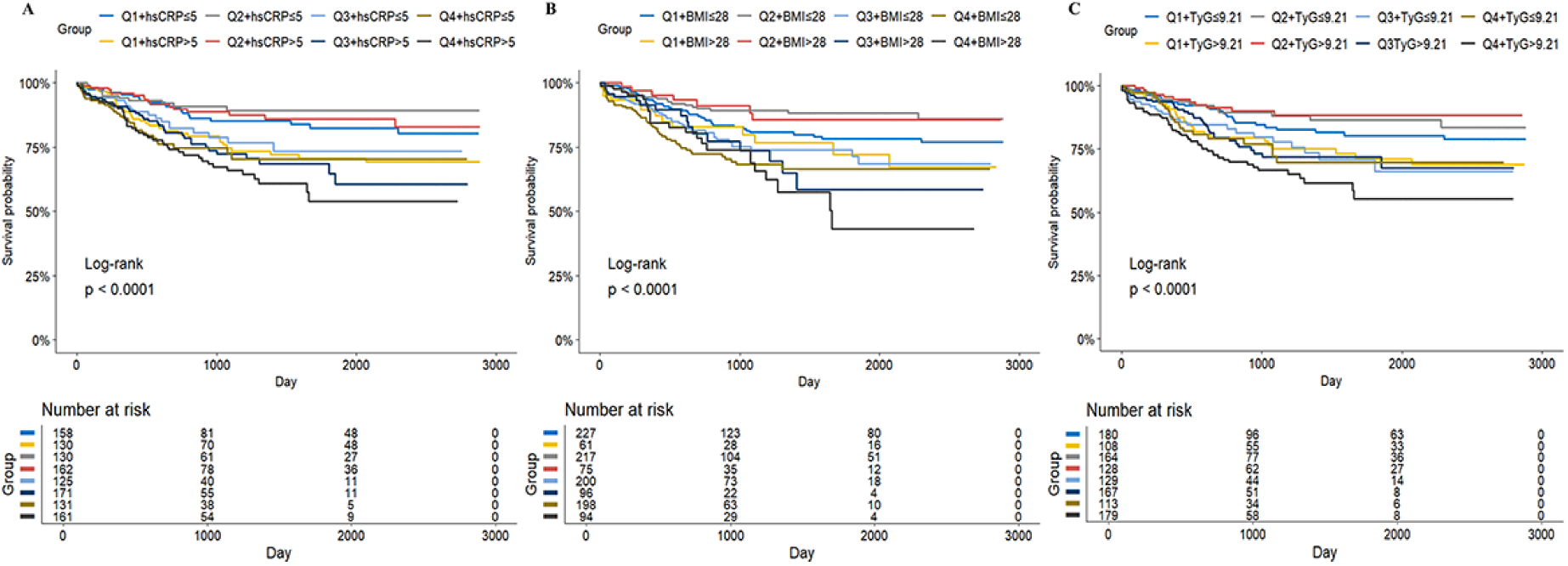
Kaplan-Meier curves for combined analysis of FFA and other indicators in patients with PMI. Kaplan-Meier curves for the occurrence of MACE in PMI patients grouped according to FFA levels combined with inflammation, obesity, and insulin resistance. **A** FFA combined with hypersensitive C-reactive protein (hs-CRP); **B** FFA combined with BMI; **C** FFA combined with TyG index.

**Table 3:**
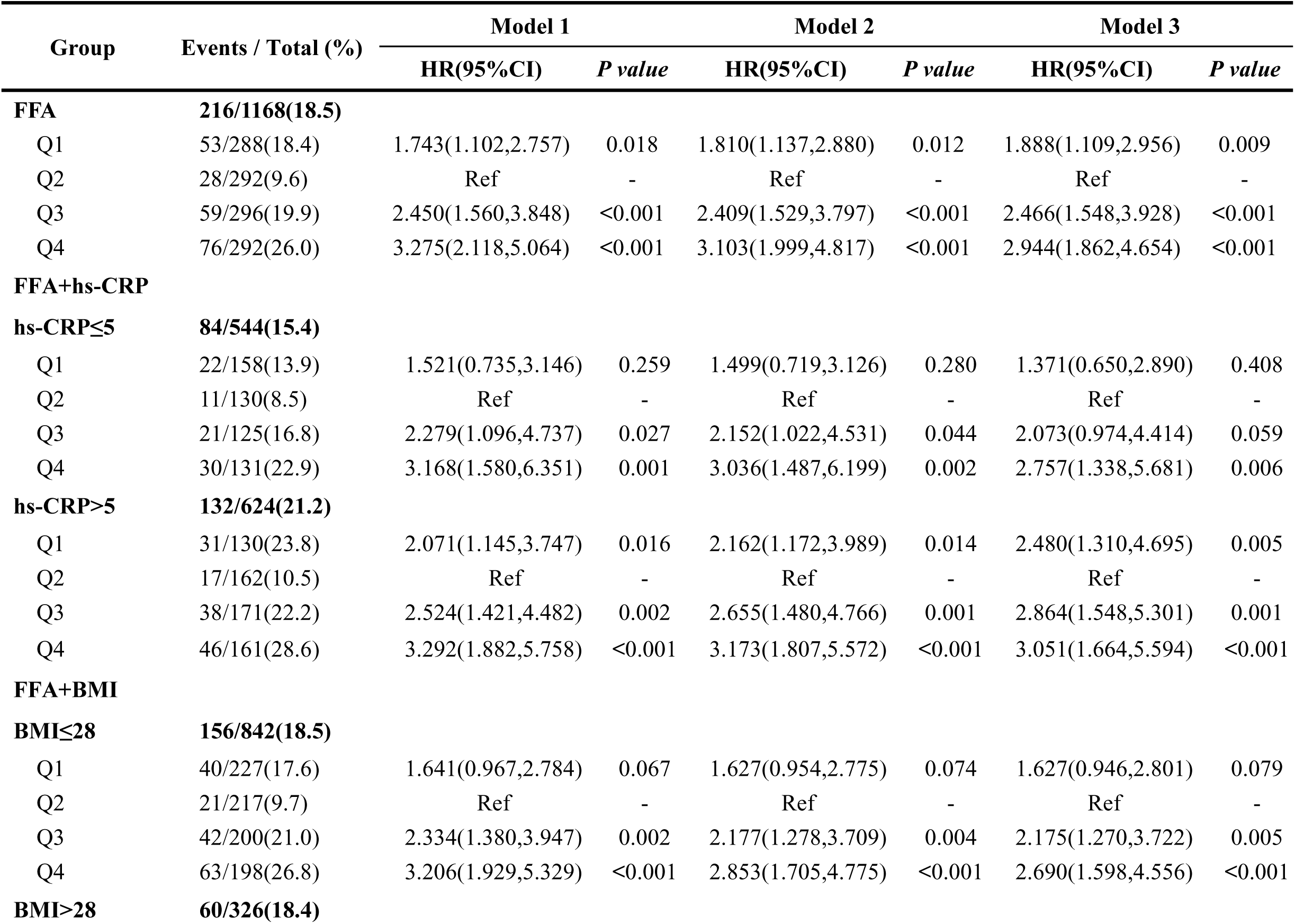
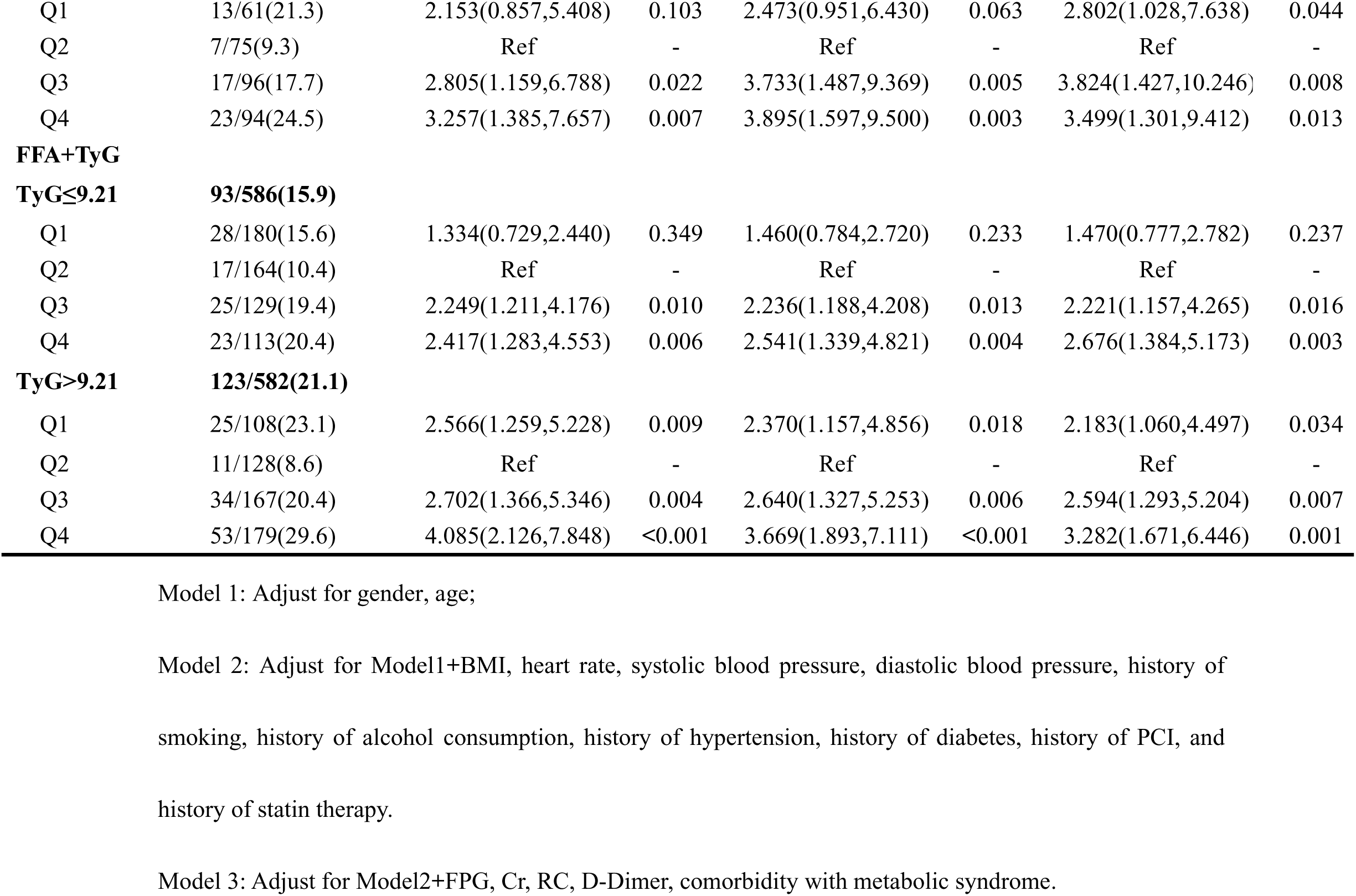
Cox proportional hazards modeling of joint role of FFA and other indicators on MACE risk.

### Sensitive analysis

To evaluate the robustness of the primary findings, a series of sensitivity analyses were performed: a. Exclusion of participants lost to follow-up (N=98) to address potential attrition bias, extreme missing assumption (MNAR) confirms the robustness of the findings; b. Re-estimation of multivariable models with additional adjustment for key clinical comorbidities, including hypertension, diabetes mellitus, and metabolic syndrome, the effect size remained stable after these adjustments; c. Stratified analyses across prespecified subgroups (sex, age, comorbidity status, and myocardial infarction type). The results of subgroup analyses suggested no significant heterogeneity between subgroups (all P for interaction>0.05); d. The impact of unmeasured confounding was quantified through E-value analysis ^17^, estimating the minimum strength of association required between hypothetical confounders and both exposure/outcome to nullify the observed hazard ratios. In the dose-effect analysis of FFA levels versus MACE, the E-values for the Q1, Q3, and Q4 subgroups were 2.301, 3.131, and 3.977, respectively. This indicates that the findings for the high-risk group would require a stronger unmeasured confounding factor to alter the interpretation of the results (see Supplementary Table 1for more details).

## Discussion

In the present study, we investigated the relationship between FFA levels and long-term MACE risk and its synergistic effect with metabolic abnormalities in PMI patients. The results showed a significant U-shaped relationship between FFA levels and MACE risk, which was particularly evident in PMI patients with comorbid MS or T2DM. Meanwhile, the joint analysis of FFA levels with high inflammatory status, obesity and insulin resistance showed that FFA levels outside a specific range exacerbated cardiovascular risk in individuals with metabolic disorders.

Relationship between FFA and prognosis confirmed in this study is consistent with the findings of several large cohort studies in recent years. In another prospective cohort study conducted in China, researchers evaluated the relationship between FFA levels and CAD and T2DM mortality in patients, and the relationship between FFA levels and risk of death showed a nonlinear U-curve, with the lowest risk at 310 µmol/L. Furthermore, at 500 µmol/L, ischemic events (MACE or MACCE) demonstrated a nonlinear U-shaped relationship, indicating the lowest risk^12^. Additionally, the team demonstrated that in patients with coronary artery disease and hypertension, FFA levels exhibited a “U-shaped” long-term prognosis. Lower FFA levels (<310 µmol/L) or higher FFA levels (≥580 µmol/L) at baseline were identified as independent risk factors for adverse outcomes^18^. However, several previous studies have shown that high levels of FFA may be an independent risk factor for cardiovascular disease^19–21^. Existing studies have mostly focused on the positive correlation between high free fatty acid (FFA) levels and poor cardiovascular prognosis. In contrast, there is still a gap in understanding the pathophysiological mechanisms by which low FFA status contributes to the long-term outcome of patients with cardiovascular disease. Based on the current state of research, we propose the following multidimensional mechanism hypothesis: Firstly, myocardial energy metabolism remodeling and contractile dysfunction^22^. More than 70% of the energy required by the normal heart is mainly provided by fatty acid oxidation, and when the level of FFA is too low, the myocardial energy supply is insufficient, leading to contractile and diastolic dysfunction; Secondly, changes in myocardial cell membrane stability and electrophysiological properties occur when FFA, a significant component of membrane phospholipids, is absent, potentially triggering malignant arrhythmias^23^; Thirdly, Imbalance in the inflammatory-oxidative stress network^24^, low FFA status may break down inherent anti-inflammatory defenses.

The mechanisms underlying the association between high FFA levels and poor prognosis in cardiovascular disease are complex and multidimensional, involving multiple pathophysiological processes such as metabolic dysregulation, inflammatory activation, oxidative stress, and organelle dysfunction. A substantial body of research has demonstrated that elevated levels of FFA can trigger vascular endothelial dysfunction through diverse mechanisms, thereby contributing to the development of atherosclerosis. For instance, FFA has been shown to induce coronary microvascular dysfunction by inhibiting the AMPK/KLF2/eNOS signaling pathway^25^; also, FFA has been shown to stabilize integrin β1 by promoting integrin β1S-nitrosylation, leading to increased heterodimerization of integrin α4β1. This, in turn, increases monocyte/macrophage adhesion to the endothelial adhesion and promotes vascular inflammation^26^. Studies of human coronary artery smooth muscle cells (HCASMC) and human coronary artery endothelial cells (hCAECs) have demonstrated that chronic FFA treatment has been shown to enhance cell-autonomous inflammatory responses through multiple mechanisms. These mechanisms include the promotion of proinflammatory gene expression and activating inflammatory signaling pathways^27,28^. Furthermore, there is a potential association between FFA and the risk of cholesterol retention and ectopic deposition within adipose tissue^29^. A Russian clinical study confirmed that elevated levels of FFA are closely associated with postinfarction myocardial remodeling and that FFA appears to be the most informative indicator of the severity of atherosclerotic coronary artery lesions^30^. Biomarker-based targeted therapeutic strategies have become the current frontier of cardiovascular research, and FFA may be potential novel therapeutic targets due to their key position in the metabolic regulatory network.

Elevated plasma levels of FFA are considered to be a common feature in populations with metabolic abnormalities. Research has revealed that a population with early-onset myocardial infarction is characterized by the presence of at least one modifiable risk factor in over 90% of cases. Among men, dyslipidemia, smoking, and substance abuse are particularly salient, while among women, diabetes mellitus, hypertension, and obesity are more prevalent^31^. In the specific group of patients with PMI, the characteristics of coronary lesions are different from those of older patients, with younger patients having a lighter atherosclerotic burden, fewer diseased vessels, shorter lesion lengths, and smaller plaque sizes^32^. Concurrently, previous studies have identified behavioral-related risk factors, including race, chronic inflammation, and persistent smoking, as significant contributors to the poor prognosis of young patients with early-onset coronary artery disease^33^. The prognostic value of FFA in metabolically dysfunctional populations shows significant heterogeneity, with predictive efficacy being particularly pronounced in subgroups of hyperinflammatory states, obesity, and insulin resistance. For instance, an analysis of the effect of FFA on the prognosis of coronary artery disease patients with different prognosis in patients with coronary artery disease with different glucose metabolic status showed that plasma FFAs were independently associated with MACE. The pre-DM+high FFAs and DM+high FFAs groups exhibited a 1.779-fold and 2.795-fold elevated risk of MACE, respectively, when compared with the NGR+low FFAs group. The findings suggest that the presence of elevated FFAs in patients with pre-DM or DM contributes to an increased risk of adverse outcomes^34^. Our findings are similar in that among those with comorbid MS or T2DM, patients with high FFA levels had a worse long-term prognosis and a greater proportion had a combined hyperinflammatory and insulin-resistant state.

In contemporary clinical research, the normal concentration range for FFA is established as 0.1–0.6 mmol/L. FFAs are vital for cardiomyocyte metabolism and play a significant role in the heart’s energy supply. Our study reveals that in the PMI population, FFA levels below 0.43 mmol/L are linked to a higher risk of MACE. This finding highlights the need for further investigation into whether low FFA levels contribute to inadequate cardiac energy supply and poor long-term outcomes for PMI patients. To tackle this issue, large-scale clinical trials are crucial to validate FFA levels as a therapeutic target and to improve the prognosis of varied metabolic populations. Developing standardized biomarker assessment and management guidelines focused on FFA is essential.

The present study had several limitations. First, the single-centre design may introduce selection bias, which needs to be validated in a multicentre cohort, although the results were shown to be robust by sensitivity analyses (E-value >2.0); Secondly, due to the relatively high daily variability of FFA levels and the close correlation between its concentration and nutritional status, fasting FFA may not adequately reflect the diurnal fluctuation of FFA. Furthermore, the relationship between the amount of change in FFA concentration during the follow-up period and the prognosis of the PMI population has not yet been explored in depth.

## Conclusions

This study is a pioneering study that reveals the correlation between different FFA levels and long-term poor prognosis in the PMI population. Due to the complex regulatory mechanisms between FFA and glycolipid metabolism and inflammatory factors, the effect of FFA on long-term poor prognosis in PMI populations is closely related to high inflammatory levels, obesity and insulin resistance status. Therefore, investigating the combined effects of FFA levels with the aforementioned metabolic disorders may provide new insights into cardiovascular and metabolic risk assessment.

## Data Availability

The datasets used and/or analysed during the current study are available from the corresponding author on reasonable request.

## Acknowledgement

We gratefully acknowledge the biobank of Tianjin Chest Hospital and the selected patients for their participation.

## Authors’ contributions

JG and YL conceived the idea and designed the study. JW made substantial contributions to data analysis, interpretation, and article writing. JXW, SR, YHW, YZ, and JXC participated in the data collection. JG, YL, CPL, and ZC participated in the drafting and critical revision of the article. All authors contributed to editorial changes in the manuscript. All authors read and approved the final version of the manuscript. All authors have participated sufficiently in the work and agreed to be accountable for all aspects of the work.

## Ethics approval and consent to participate

The study protocol was reviewed and endorsed by the Internal Review Board of Tianjin Chest Hospital (No. 2017key -007-01). Before enrollment, all patients provided written informed consent to the Declaration of Helsinki and its subsequent amendments or equivalent ethical standards.

## Consent for publication

Not applicable.

